# Development and Validation of a Multiplex Real-Time PCR Assay for Detection and Quantification of *Streptococcus pneumoniae* in Pediatric Respiratory Samples

**DOI:** 10.1101/2023.05.03.23289397

**Authors:** Molly Butler, Garrett Breazeale, Eric Mwangi, Elaine Dowell, Samuel R Dominguez, Linda Lamberth, Kristina Hultén, Sarah A Jung

## Abstract

*Streptococcus pneumoniae* (*Spn*) is a bacterial pathogen that causes a range of disease manifestations in children, from acute otitis media to pneumonia, septicemia, and meningitis. Primary *Spn* laboratory diagnostic identification methods include culture, antigen testing, single-plex real-time PCR, and syndromic PCR panels. However, each method lacks sensitivity, specificity, and/or cost efficiency. We developed and validated a quantitative, multiplex PCR assay that uses three *Spn* genomic targets (*lytA, piaB*, and SP2020) for improved sensitivity and specificity to detect *Spn* in pleural fluid (PF), bronchoalveolar lavage (BAL), tracheal aspirate (TA), and upper respiratory (UR, research only) samples. Validation testing included analytical sensitivity (limit of detection), specimen storage, analytical specificity (cross-reactivity), and accuracy studies. Limit of detection is 500 genome copies/mL in lower respiratory samples and 100 copies/mL in upper respiratory specimens, with quantification range of 1,000 to 10,000,000 copies/mL. Specimens can be stored frozen at least 60 days and *Spn* DNA is stable through 3 freeze-thaw cycles. No cross-reactivity was observed against 20 closely related microorganisms and/or microorganisms that can be detected in similar sample types, including *Streptococcus pseudopneumoniae*. In reference range testing, *Spn* was detected in 5 of 23 (21.7%) PF, 2 of 19 (10.5%) BAL, 1 of 20 (5.0%) TA, and 44 of 178 (24.7%) UR residual specimens. For accuracy studies, 98 specimens were tested and overall percent agreement with a qualitative, *lytA*-based comparator assay was 96.9% across all sample types. This multiplex, quantitative PCR assay is a sensitive and specific method for *Spn* detection in pediatric respiratory samples.

## Introduction

*Streptococcus pneumoniae* (*Spn*) is a significant bacterial pathogen. While non-pathogenic colonization of the upper respiratory tract is common, *Spn* can cause a range of disease states including acute otitis media, pneumonia, bacteremia, and meningitis (1-3). As of 2016, *Spn* was the leading cause of lower respiratory tract infection (LRTI) morbidity and mortality worldwide (1, 2, 4). When compared with other common pathogens associated with LRTI (*Haemophilus influenzae*, influenza virus, and respiratory syncytial virus), *Spn* was found to cause more deaths than these three other pathogens combined (4).

A necessary prerequisite for invasive pneumococcal disease (IPD) is *Spn* upper respiratory colonization (1). Rates of nasopharyngeal (NP) carriage of *Spn* are highest in children aged 2 to 3 years, reaching near or above 50%, although exact colonization rates vary by geographic location, community vaccination status, and socioeconomic factors (1, 5). Children less-than 5 years have highest *Spn* colonization rates and have highest *Spn*-associated morbidity and mortality compared to other age groups (4).

Accurate *Spn* laboratory identification from lower respiratory sources during confirmed or suspected LRTI can aid in clinical management and initiation of appropriate antimicrobial therapy. Current commercial and research *Spn* identification methods include standard microbiology culture techniques, antigen testing, real-time PCR, and syndromic multiplex PCR panels. Each of these methods has certain disadvantages. The viability of *Spn* to grow in culture deteriorates in standard laboratory storage conditions due to quorum sensing regulated-autolysis and is frequently hindered by antibiotic pretreatment (6-8). If culture growth occurs, phenotypic identification techniques, including optochin susceptibility and bile solubility, are characteristics shared by other members of the *Streptococcus mitis* group, and some *Spn* isolates may exhibit optochin resistance or bile insolubility (9). Antigen testing, most commonly performed on urine specimens, has unclear clinical utility in diagnosing pneumonia in children due to relatively high *Spn* colonization rates (10). Syndromic PCR panels are valuable tools for rapid diagnosis, but lack cost effectiveness, particularly if suspicion is high for a specific pathogen and are often restricted to only the sample types that have received U.S. Food and Drug Administration approval.

Distinction between *Spn* and closely related species, particularly *Streptococcus pseudopneumoniae*, by single-plex real-time PCR is challenging due to the high degree of genetic transfer between and within *Spn* isolates, closely related bacterial species, and co-colonizing organisms of the upper respiratory tract (9, 11). *Spn* gene targets commonly used in PCR include *lytA*–which has been identified in several *Streptococcus mitis* isolates–and *piaB*, which is not always encoded by *Spn* isolates (9, 12-15). Recent work to identify genomic markers universally and exclusively found within *Spn* isolates identified SP2020, a genomic region encoding a putative transcriptional regulator (14, 15).

Given the high degree of genetic diversity within *Spn* isolates and the potential for any given single gene target to be found in a non-*Spn* species, we designed a multiplex real-time PCR which targets three *Spn* genomic regions (*lytA, piaB*, and SP2020) for improved sensitivity and specificity to detect *Spn* in respiratory samples. In this study, we evaluated the performance of this assay in lower respiratory specimens including pleural fluid (PF), bronchoalveolar lavage (BAL), and tracheal aspirate (TA) samples for clinical purposes, as well as in upper respiratory specimens collected in saline, skim milk tryptone glucose glycerol (STGG, media type used for *Spn* recovery (16)), or viral/universal transport media (VTM/UTM) for research purposes.

## Methods

### Center and Laboratory Description

This study was conducted in the Clinical Microbiology Laboratory at Children’s Hospital Colorado (CHCO). CHCO is a quaternary academic pediatric hospital with greater-than 500 beds serving the Denver metropolitan area, the state of Colorado, and seven surrounding states. Residual specimens used in this study were samples previously collected, tested, and reported for clinical and research purposes. Study investigators conducted retrospective chart review from the electronic health record. Approval was obtained from the Colorado Multiple Institutional Review Board (COMIRB # 21-4052).

### Reference and Clinical Material

The reference strain used in this study was *Streptococcus pneumoniae* serotype 19F (ATCC® 49619). Quantified whole organism *Spn* (strain 19F, reference number 0801439) used for limit of detection (LOD) studies and construction of standard curve was obtained from ZeptoMetrix (Buffalo, NY, USA). The titer of this material was determined by culture techniques, thus stock concentration reported by the manufacturer is in colony forming units per milliliter (CFU/mL). However, a more appropriate unit for reporting concentration by nucleic acid amplification assays is genome copies per mL (copies/mL). Therefore, we relied on the hypothesized equivalency that 1 CFU = 1 genome and assumed all specimens used for LOD and standard curve construction were equivalent to clinical and contrived specimens used for analytical testing.

Clinical *Spn* isolates used in this study were isolated in the CHCO Clinical Microbiology Laboratory from BAL and blood cultures. Isolates grown from BAL fluid were identified as *Spn* by Gram stain, catalase testing, hemolysis characterization, and optochin sensitivity. Isolates grown in blood culture were identified as *Spn* by the BioFire^®^ FilmArray Blood Culture Identification Panel (BCID) or Blood Culture Identification Panel 2 (BCID2, BioFire Diagnostics, Salt Lake City, UT, USA). Clinical samples used for LOD, reference range, and accuracy studies for lower respiratory sample types were residual PFs, BALs, and TAs previously collected for routine clinical testing. Clinical samples used for reference range and accuracy studies from upper respiratory sample types were residual NP swab samples in saline or UTM/VTM previously tested on BioFire^®^ Respiratory Pathogen Panel 2.1 test (RPP, BioFire Diagnostics, Salt Lake City, UT, USA). STGG was prepared in-house and composed of skim milk (2% w/v), tryptic soy broth (3% w/v), glucose (0.5% w/v), and glycerol (10% v/v) and sterilized by autoclave.

Non-pneumococcal strains used for analytical specificity studies included *Streptococcus mitis* (ATCC^®^ 49456), *Streptococcus salivarius* (ATCC^®^ 25975), *Streptococcus pyogenes* (ATCC^®^ 19615), *Streptococcus agalactiae* (ATCC^®^ 12386), *Staphylococcus aureus* (ATCC^®^ 29213), *Staphylococcus epidermidis* (ATCC^®^ 12228), *Legionella pneumophila* (ATCC^®^ 33152), *Haemophilus influenzae* (ATCC^®^ 79247), *Pseudomonas aeruginosa* (ATCC^®^ 27853), *Aerococcus viridans* (ATCC^®^ 11563). For cross-reactivity studies against *Streptococcus. pseudopneumoniae*, solutions in sterile water were prepared from plated media of ATCC^®^ strain BAA-960 ranging in concentration from 0.5 to 2.0 McFarland standard. The NATtrol Respiratory Verification Panel 2 (ZeptoMetrix, Buffalo, NY) was used for cross-reactivity studies of molecular material against *Mycoplasma pneumoniae, Chlamydophila pneumoniae*, influenza A virus (A/NewCal/20/99, A/Brisbane/10/07, A/NY/02/11), influenza B virus (B/Florida/02/06), rhinovirus, and respiratory syncytial virus A. SARS-CoV-2 cross-reactivity was evaluated by using known SARS-CoV-2-positive samples and Exact Diagnostics SARS-CoV-2 standard control material (Exact Diagnostics, Fort Worth, TX, USA).

### PCR Protocol

Nucleic acid, including internal positive control DNA (ThermoFisher, Waltham, MA, USA), was isolated on Qiagen EZ1 Advanced XL platform using the DNA Tissue Kit (Qiagen, Hilden, Germany) and subjected to real-time PCR analysis on an ABI 7500 real-time PCR system (ThermoFisher, Waltham, MA, USA). PCR reaction components included PerfeCTa qPCR Tough Mix, Low ROX (Quantabio, Beverly, MA, USA), forward primer, reverse primer, and probe for each of the three *Spn* targets at final reaction concentration of 200 nM, internal positive control primers and probe mix (ThermoFisher, Waltham, MA, USA), and molecular grade water for a 25 μL reaction volume. Primer and probe sequences are listed in **Table 1**. Real-time PCR cycling conditions were 95°C x 10 minutes, followed by 45 cycles of 95°C x 15 seconds and 60°C x 1 minute.

**Table 1:**
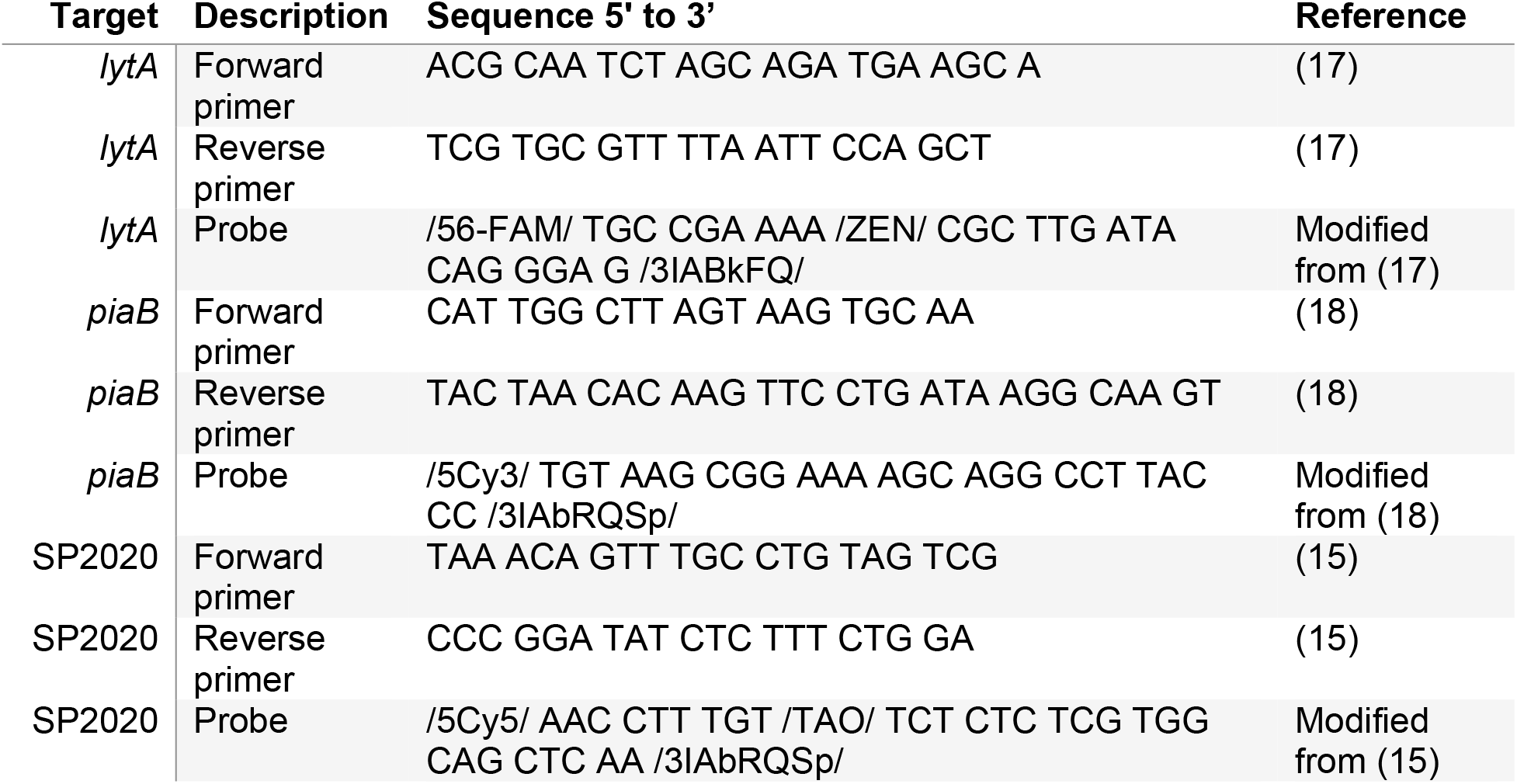
Primer and probe sequences

### Interpretation Criteria

A C_T_ < 40 was considered positive amplification for each *Spn* target (*lytA, piaB*, and SP2020). A sample was then considered *Spn*-positive if at least 2 of the 3 targets were detected with C_T_ < 40. A sample was considered negative if 0-1 of the 3 *Spn* targets were detected with C_T_ < 40, and the internal positive control C_T_ value was < 35. A sample was considered invalid if there was no amplification of any *Spn* targets, and the internal positive control C_T_ value was ≥ 35.

### Accuracy

The comparator assay used for accuracy studies is a *lytA-*targeting qualitative real-time PCR for *Spn* molecular detection. Quantification of positive samples sent for accuracy studies ranged from less-than 1,000 copies/mL to greater-than 10,000,000 copies/mL. Samples sent included both clinical and contrived specimens in lower respiratory matrices (PF) and upper respiratory matrices (saline and STGG). A summary of sample type distribution is presented in **Table 2**.

**Table 2:**
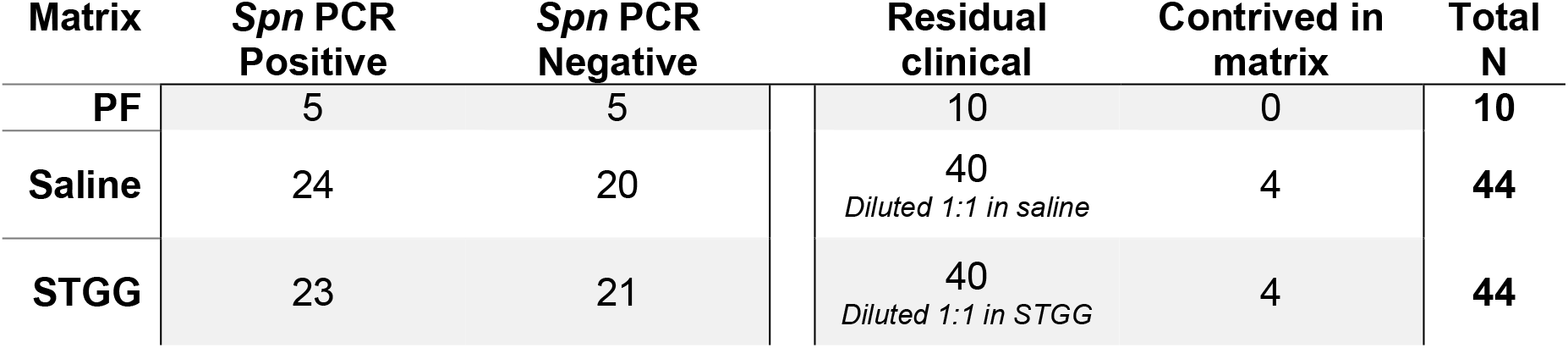
Samples sent for accuracy studies. A total of 98 samples were sent for testing, representing both clinical and contrived specimens. Quantification range of *Spn* PCR positive samples was <1,000 copies/mL to >10,000,000 copies/mL.

### Statistics

Statistical analyses, including linear regression and coefficient of determination, were completed in R version 4.0.3.

## Results

### Analytical Sensitivity

To determine analytical sensitivity, we prepared two independent dilution series of *Spn* in each matrix (PF, BAL, TA, saline, STGG, and VTM), ranging from 10^7^ to 10^1^ copies/mL. At least 10 replicates at each concentration were tested over two consecutive days. To establish interpretation criteria, we evaluated the relationship between each of three targets over a range of concentrations from 10^7^ through 10^2^ copies/mL (**Figure 1A-C**). *piaB* had the lowest efficiency of the three targets, but there was a high degree of correlation (R^2^ > 0.9) between the C_T_ values of each of the three targets when compared pairwise across the range of concentrations (**Figure 1A-C**). At the lowest concentration (100 copies/mL), we observed consistent amplification of *lytA* and SP2020 with inconsistent *piaB* amplification. We therefore established positivity criteria to be amplification of at least two targets before 40 cycles.

**Figure 1:**
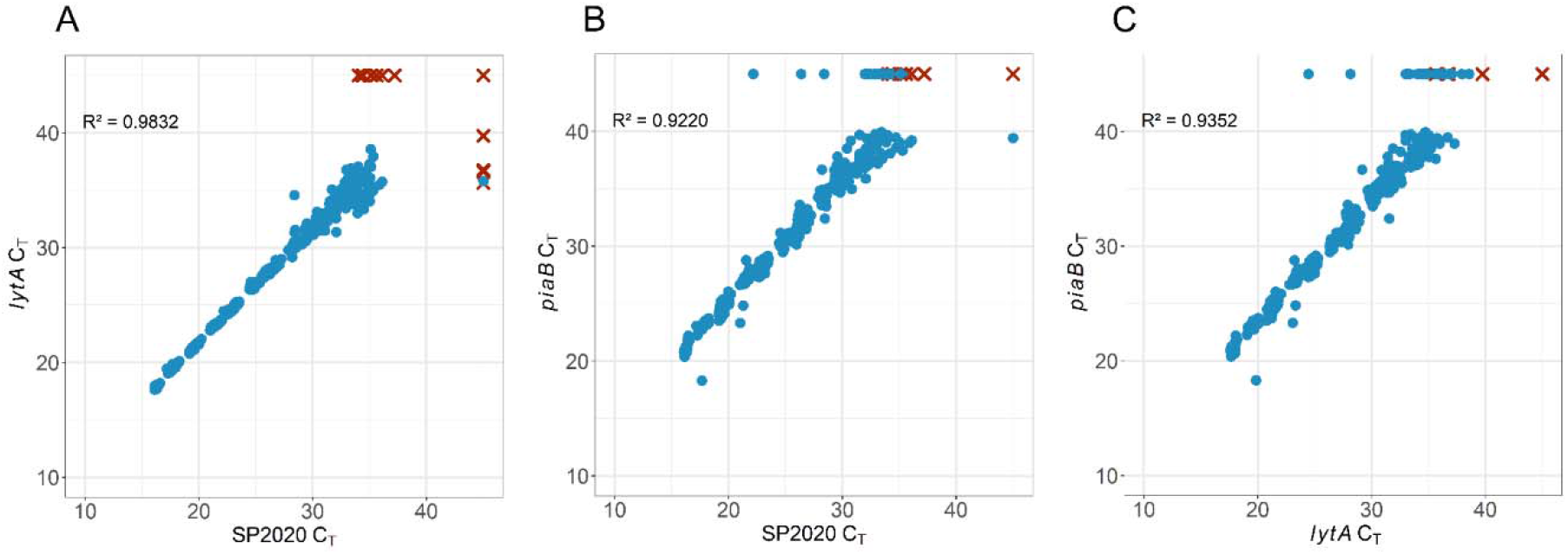
Pairwise comparison of C_T_ values of *lytA*, SP2020, and *piaB* over the range of 10^7^ to 10^2^ *Spn* copies/mL. At least 10 replicates were tested at each concentration in each matrix (PF, BAL, TA, saline, STGG, and VTM). Replicates meeting detection criteria (at least two targets with C_T_ < 40) indicated by blue circles. Replicates not meeting detection criteria indicated by red X marks. No amplification indicated as C_T_ = 45. R^2^ as indicated for each pairwise comparison calculated from replicates meeting detection criteria only.

We found 100% *Spn* detection in all sample types at 1,000 copies/mL. For lower respiratory matrices (PF, BAL, and TA), *Spn* was detected in 100% of replicates at 500 copies/mL, but we observed less reliable detection at 100 copies/mL (< 90%). For upper respiratory matrices (saline, VTM, and STGG), we observed 100% detection at 100 copies/mL but minimal detection at 10 copies/mL (< 10%). Therefore, we established the LOD to be 500 copies/mL in lower respiratory sample types and 100 copies/mL in upper respiratory sample types (**Table 3**).

**Table 3:** Percentage of replicates (n = at least 10) in each matrix that met *Spn* detection criteria (at least two targets with C_T_ < 40). NT indicates not tested.

|  | Copies/mL |  |  |  |
| --- | --- | --- | --- | --- |
|  | 1,000 | 500 | 100 | 10 |
| **PF** | 100% | 100% | 83.3% | 8.3% |
| **BAL** | 100% | 100% | 77.8% | 0% |
| **TA** | 100% | 100% | 44.4% | 0% |
| **Saline** | 100% | NT | 100% | 8.3% |
| **STGG** | 100% | NT | 100% | 0% |
| **VTM** | 100% | NT | 100% | NT |

Due to the reliably lower C_T_ values of SP2020 compared to the other two targets at each tested concentration, we elected to use SP2020 C_T_ values for construction of a standard curve to determine *Spn* quantification. Comparison of polynomial and linear regression revealed a loss of linearity at the highest concentration tested (1 × 10^8^ copies/mL, data not shown), which led us to establish an upper limit of quantification (LOQ) at 1 × 10^7^ copies/mL. We selected a lower LOQ of 1 × 10^3^ copies/mL because it was the lowest concentration tested at which the standard deviation of SP2020 C_T_ values was less-than 1 C_T_ value. Standard curves constructed in each of the matrices analyzed were consistent, with a y-intercept range from 39.9-41.9 and a slope range from -3.6 to -3.3 (**Figure 2**). Lower respiratory matrices (PF, BAL, and TA) had consistently higher C_T_ values across the concentration range, likely due to the presence of PCR inhibitors in these sample types.

**Figure 2:**
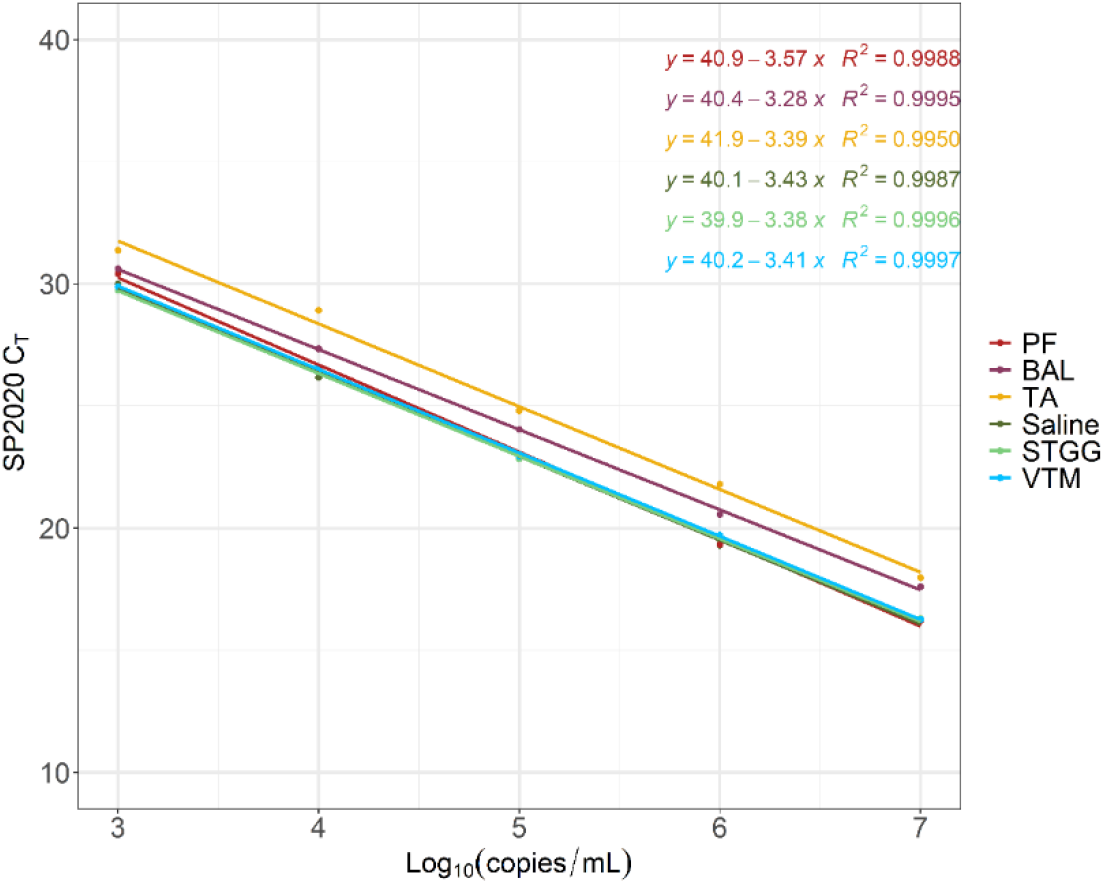
Average SP2020 C_T_ values for each tested concentration of standard for each matrix with indicated standard curve and R^2^ value. N = at least 10 replicates at each concentration in each matrix.

We also assessed assay sensitivity against *Spn* clinical isolates. These clinical isolates were previously identified by our laboratory as *Spn* by standard microbiology culture techniques (n = 8) or by BCID/BCID2 (n = 9). Of these, 16/17 (94.1 %) were *Spn*-positive by PCR. The single isolate that was identified by standard culture techniques to be *Spn* but was PCR-negative was sent to an independent, reference laboratory and was identified as “*S. mitis*-group, not *S. pneumoniae*.

### Specimen Storage and Precision

To evaluate acceptable specimen storage conditions, stocks of high-, medium-, and low-positive and negative samples in saline and STGG were stored at room temperature, 4°C, - 20°C and -70°C. High-positive samples were defined as those greater-than 100,000 *Spn* copies/mL, medium-positive defined as *Spn* 1,000-20,000 copies/mL, and low-positive defined as less-than *Spn* 1,000 copies/mL (below established LOQ). Stocks at room temperature were tested at 24 hours and 48 hours; stocks at 4°C were tested at 4 days, 7 days, and 14 days; stocks at -20°C were tested at 10 days, 21 days, and 30 days; and stocks at -70°C were tested at 10 days, 21 days, 30 days, and 60 days. Stocks tested at frozen temperatures underwent a freeze-thaw cycle at each tested time point. Overall standard deviation for each gene target C_T_ value was less-than 1.5 C_T_ (data not shown) and overall standard deviation for quantification was within 0.5 log_10_(copies/mL) across all storage temperatures and duration (**Figure 3**). However, there was a notable increase in C_T_ values for all gene targets after a fourth freeze-thaw cycle (samples stored at -70°C for 60 days), corresponding to a reduction in calculated copies/mL (**Figure 3**, darkest blue “+” symbols). These data suggest that samples should not be subjected to more than three freeze-thaw cycles to maintain quantification accuracy.

**Figure 3:**
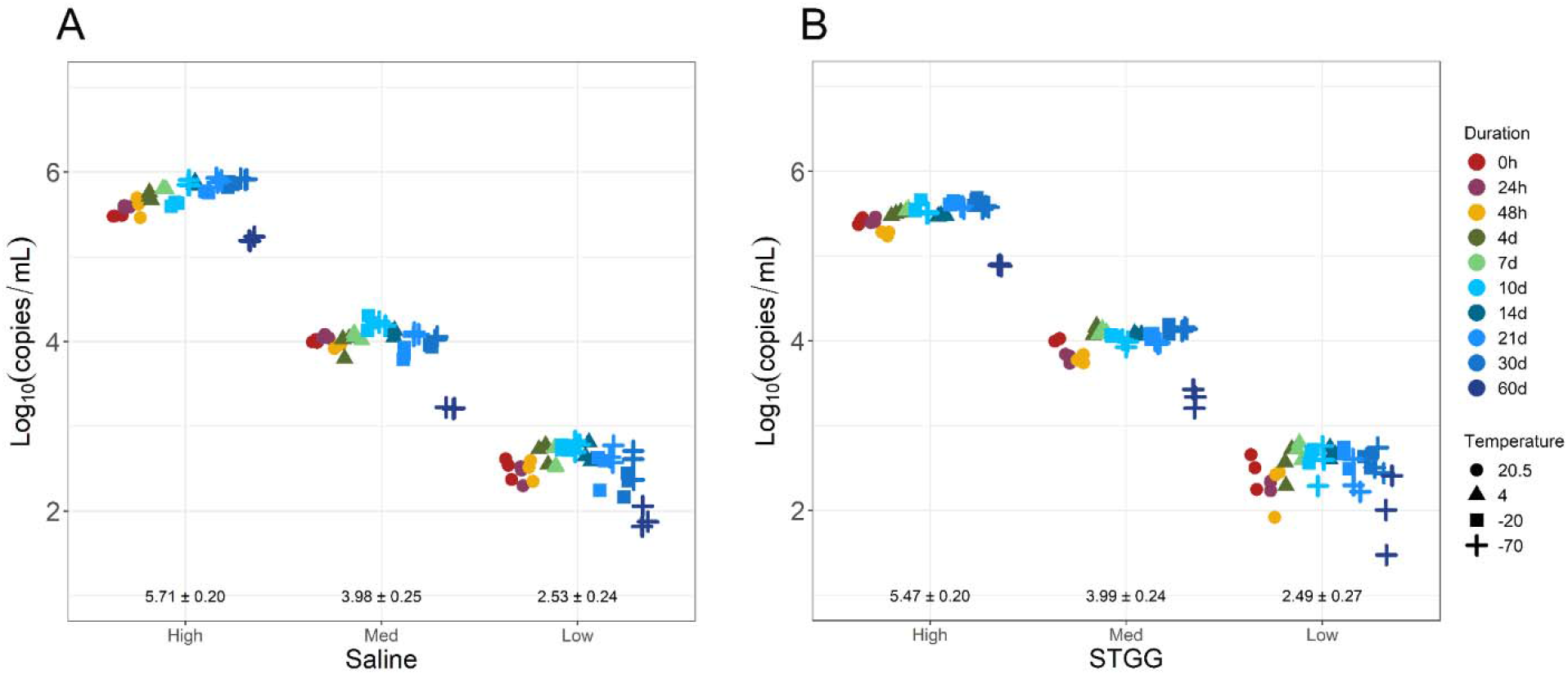
Calculated *Spn* quantification in high-, medium-, and low-positive samples prepared in saline (**A**) or STGG (**B**). Storage duration indicated by color; storage temperature in °C indicated by shape. Each sample at each storage condition was tested in triplicate. Mean ± SD indicated for each sample.

For reproducibility studies, high-, medium-, and low-positive control samples, as well as negative controls, prepared in saline or STGG were independently extracted and analyzed by real-time PCR over six days by three independent technologists. Variability in C_T_ value was the highest among low-positive samples. However, in both matrices overall standard deviation was less-than 1 C_T_ value (data not shown) and within 0.5 log_10_(copies/mL) (**Figure 4**).

**Figure 4:**
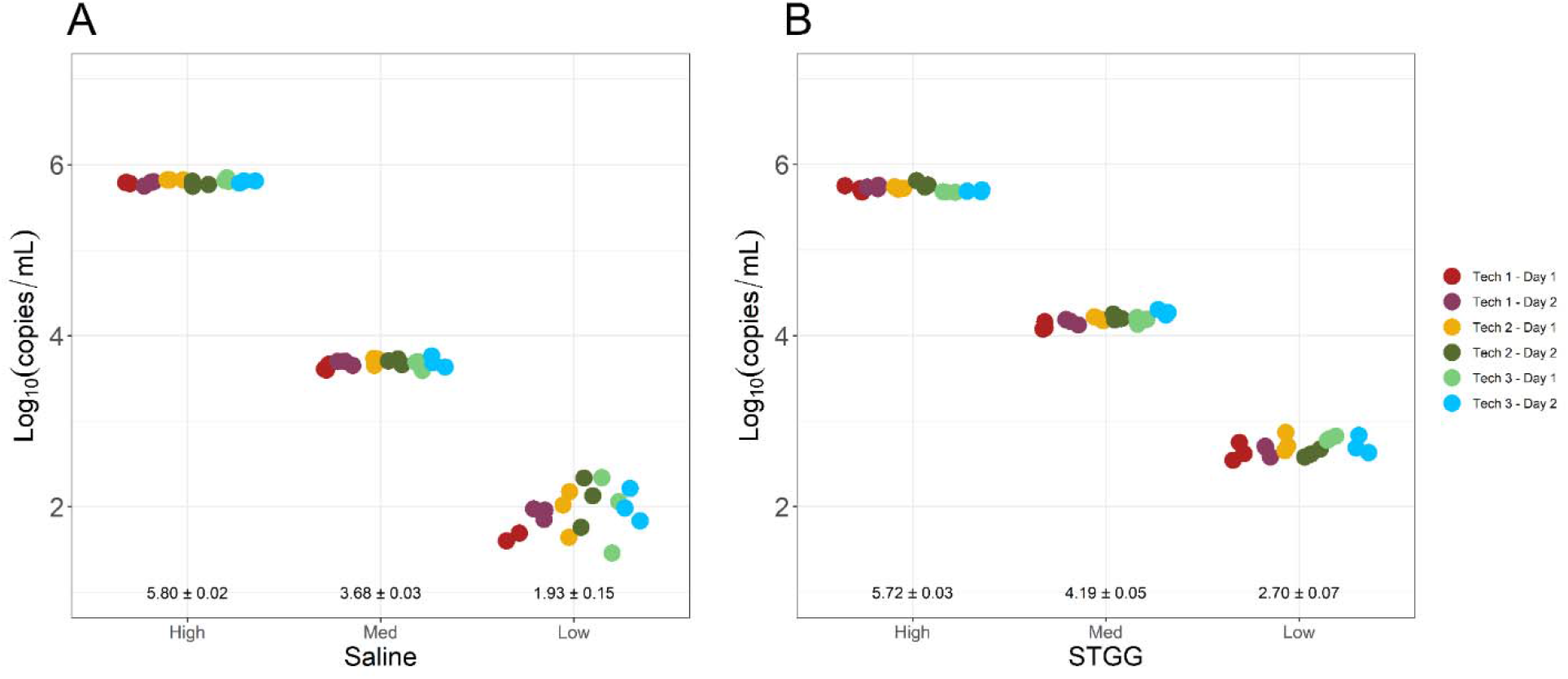
Calculated *Spn* quantification in high-, medium-, and low-positive control samples prepared in saline (**A**) or STGG (**B**). Each independent run indicated by a unique color. Samples during each run were tested in triplicate. Mean ± SD indicated for each sample.

### Analytical Specificity

We performed cross-reactivity studies against organisms genetically related to *Spn*, as well as organisms likely to be found in similar sample types. Tested three times each, we observed no cross-reactivity with the following streptococcal species: *S. mitis, S. salivarius, S. pyogenes*, and *S. agalactiae*. Also tested three times each, we observed no cross-reactivity to *Staphylococcus aureus, Staphylococcus epidermidis, Legionella pneumophila, Haemophilus influenzae, Pseudomonas aeruginosa, Aerococcus viridans, Mycoplasma pneumoniae, Chlamydophila pneumoniae*, influenza A virus, influenza B virus, rhinovirus, respiratory syncytial virus A, and SARS-CoV-2.

Due to the high genetic similarity between *Spn* and *Streptococcus pseudopneumoniae*, we performed a more extensive cross-reactivity investigation between these two streptococcal species. We tested a range of *S. pseudopneumoniae* concentrations over multiple days for a total of 30 replicates. We observed limited and sporadic (not associated with concentration) single-target amplification with *S. pseudopneumoniae*. Late amplification of the *lytA* target occurred in 6 of 30 replicates (20.0%), with C_T_ values greater-than 40. Positive amplification was also observed with the SP2020 target in 1 of 30 replicates (3.3%), with a C_T_ value less-than 40 (C_T_ = 36.4). There was no detectable amplification of the *piaB* target in any of the 30 replicates. None of the 30 replicates would have met *Spn* detection criteria.

### Reference Range

To establish a reference range for detecting *Spn* in lower respiratory samples, we tested 23 residual PF samples collected over 2 years and previously tested by routine clinical culture. *Spn* was detected by PCR in 5 of 23 samples (21.7%). *Spn* was identified by PF culture in 1 of the 5 PCR-positive specimens. We tested 19 residual BAL samples and 20 TAs collected over 2 months that had been previously tested by routine clinical culture, RPP testing, and/or FluVID testing (SARS-CoV-2, influenza A virus, influenza B virus, respiratory syncytial virus). *Spn* was detected by PCR in 2 of 19 residual BALs (10.5%) and in 1 of 20 TAs (5.0%). *Spn* was identified through routine culture in both *Spn* PCR-positive BAL specimens but not in the *Spn* PCR-positive TA. In each of the three lower respiratory matrices (PF, BAL, TA), we observed at least one instance of PCR inhibition, with 4 of 62 (6.5%) specimens resulted as “Invalid.” The comparison between *Spn* PCR and culture in lower respiratory specimens is summarized in **Table 4**.

**Table 4:** Comparison between *Spn* PCR and standard culture of residual lower respiratory specimens (n = 22 PFs, 15 BALs, and 12 TAs). Specimens which were not previously tested by routine culture were excluded (n = 2 BALs, 7 TAs); specimens which resulted as “Invalid” were excluded (n = 1 PF, 2 BALs, 1 TA).

|  | ***Spn* Culture**<br>**Positive** | ***Spn* Culture**<br>**Negative** |
| --- | --- | --- |
| ***Spn* PCR**<br>**Positive** | 3 | 5 |
| ***Spn* PCR**<br>**Negative** | 0 | 41 |

Additionally, we tested 178 residual NP swab specimens in saline or universal transport media (UTM) that had been previously tested by RPP. Samples were collected over 6 months from patients 5 years or younger. *Spn* was PCR-detected in 44 of 178 specimens (24.7%). Reference range results are summarized in **Table 5**.

**Table 5:** Reference range results for PF, BAL, and TA, and upper respiratory (NP swab) sample types.

|  | Total Tested<br>N | *Spn* Detected<br>N (%) |
| --- | --- | --- |
| PF | 23 | 5 (21.7) |
| BAL | 19 | 2 (10.5) |
| TA | 20 | 1 (5.0) |
| NP Swab | 178 | 44 (24.7) |

### Accuracy

Samples representing a range of high-, medium-, and low-positive results, as well as negative samples, were sent to an independent laboratory for accuracy studies. The comparator assay is a *lytA*-based, qualitative PCR for detection of *Spn*. Positive percent agreement among PF samples was 83.3% and percent negative agreement was 100%. The single discordant PF sample (*Spn* PCR negative but comparator assay positive) had an unexpectedly high C_T_ value for the internal positive control, indicative of PCR inhibition. For saline samples, there was both 100% negative and positive percent agreement in qualitative interpretation. STGG samples also had a 100% positive agreement. Two STGG samples classified as low-positive (less-than 1,000 copies/mL) by our *Spn* PCR were classified as negative by the comparator assay, yielding a 91.3% negative agreement. Positive percent agreement across all specimen sources was 98.0%, negative percent agreement was 95.7%, and overall percent agreement was 96.9% (**Table 6)**.

**Table 6:** Qualitative agreement between this multiplex *Spn* PCR and comparator, *lytA*-based PCR.

|  | Comparator Positive | Comparator Negative |
| --- | --- | --- |
| *Spn* PCR Positive | 50 | 2 |
| *Spn* PCR Negative | 1 | 45 |

Though an imperfect comparison due to the multiplex nature of our assay, we found an acceptable degree of correlation between our *lytA* C_T_ values and the *lytA* C_T_ values produced by the comparator assay (PF R^2^ = 0.91, saline R^2^ = 0.77, STGG R^2^ = 0.73) (**Figure 5**).

**Figure 5:**
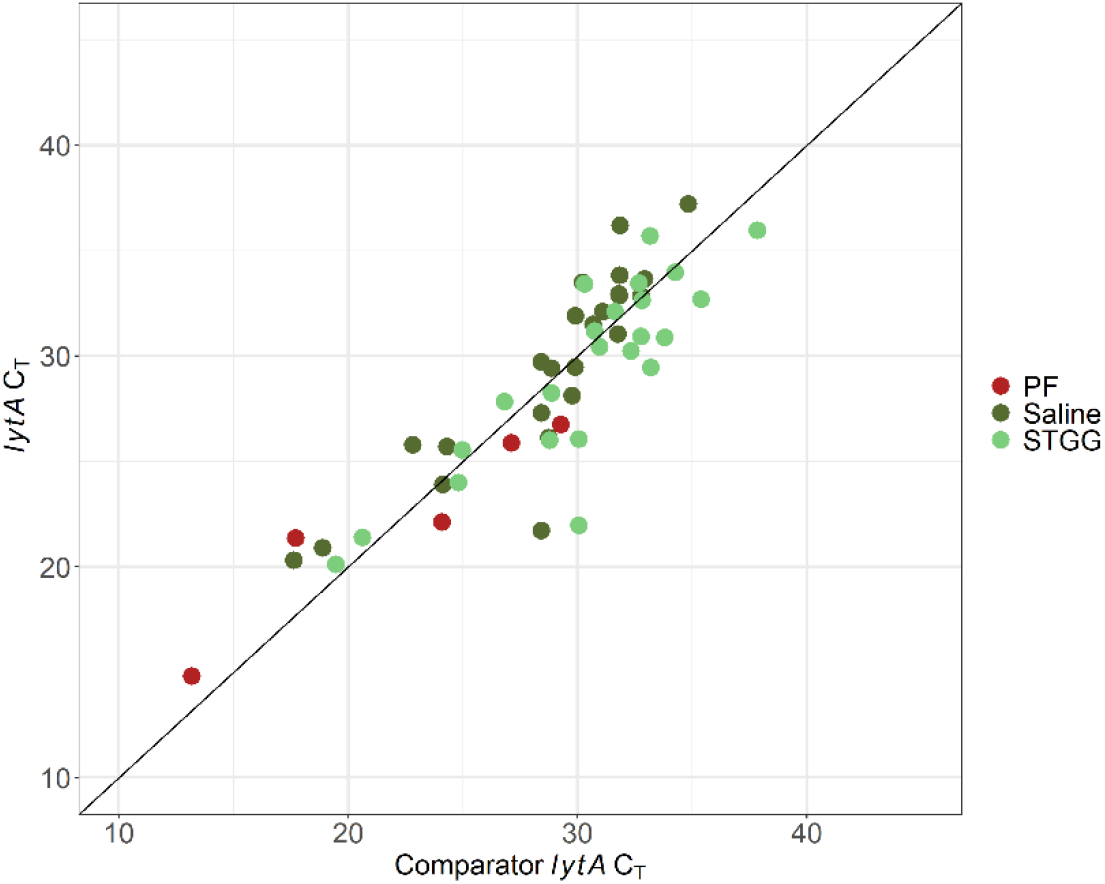
Comparison of *lytA* C_T_ values from the comparator assay and this assay. *y = x* indicated by black line.

## Discussion

Accurate and reliable *Spn* detection is essential for appropriate disease management in lower respiratory sources and for understanding colonization dynamics in upper respiratory sources. Despite this need, traditional *Spn* microbiological methods (primarily culture and antigen testing), as well as current PCR methods, lack sensitivity and specificity due to both high genetic diversity within and between *Spn* strains and characteristic phenotypic and genotypic *Spn* elements in non-*Spn* species (9, 12, 15, 19).

To address this need, we developed and validated a quantitative real-time PCR targeting three *Spn* genomic regions for improved sensitivity and specificity in *Spn* detection in respiratory specimens. This assay detects as few as 500 *Spn* genome copies/mL and quantification was linear across 6 orders of magnitude. Quantification remained consistent after storage in typical laboratory conditions and through three freeze-thaw cycles. Cross-reactivity was not observed against any organism tested.

We evaluated the assay’s performance in PF, BAL, and TA specimens as well as in upper respiratory specimens (NP swab samples collected in saline, UTM/VTM, or STGG). The validation of this assay in both lower and upper respiratory specimens allows its use to address clinical and research questions. In clinical contexts, accurate and timely *Spn* identification in lower respiratory specimens is important for initiation of appropriate antimicrobial therapy and medical management of pneumococcal disease. Using our assay, we detected *Spn* in lower respiratory samples from which *Spn* was not identified through routine laboratory testing. Due to the success of this validation study, *Spn* PCR from PF, BAL, and TA specimens is now available for clinical use in our laboratory. Quantification of *Spn* from lower respiratory specimens is reported in semi-quantitative ranges due to consistent elevation of SP2020 C_T_ values across the quantifiable range (**Figure 2**). The semi-quantitative ranges that we use for clinical reporting are: less-than 1,000 copies/mL, 1,000-10,000 copies/mL, 10,001-100,000 copies/mL, 100,001-1,000,000 copies/mL, and greater-than 1,000,000 copies/mL.

For research purposes, this *Spn* PCR may be used for understanding *Spn* colonization dynamics in the upper respiratory tract. Understanding these dynamics is an area of ongoing research since *Spn* colonization is a necessary prerequisite for *Spn* disease. Additionally, there are several described relationships between *Spn* colonization and viral respiratory disease, which have important implications for invasive disease prevention (1, 5, 20-26). Application of our assay may include characterizing *Spn* detection and bacterial load in the upper respiratory tract and association with viral co-infection and incidence of invasive pneumococcal disease.

This study has several notable limitations. First, this study was conducted at a single institution and is therefore restricted to *Spn* strains available commercially (ATCC^®^) or which are currently circulating in the Denver, CO geographic area. Additionally, this study was limited to a pediatric patient population, and there are known age-related differences in *Spn* serotype prevalence (27). This study was conducted during widespread circulation of SARS-CoV-2, which has unknown implications on *Spn* colonization and disease dynamics (20). As with all nucleic acid amplification tests, the assay’s sensitivity and specificity may be affected by genetic variation in the target organism and will be routinely evaluated against currently circulating strains. The inclusion of three *Spn* gene targets in this assay should aid in the early identification of such issues if single-target drop-out is observed with increasing frequency. In such a case, primer-probe sequences may be updated to maintain assay performance of this assay. In conclusion, this laboratory-developed test showed robustness against all studies performed. The performance of this assay is acceptable for clinical and research purposes from pediatric lower and upper respiratory sources, respectively.

## Data Availability

All data produced in the present study are available upon reasonable request to the authors.

## Acknowledgements and Disclosures

Most of the funding and resources were provided by the Department of Laboratory Medicine and Pathology at Children’s Hospital Colorado. A portion of the funding and resources was provided by Pfizer (New York, NY) for a separate, collaborative project, of which the data and conclusions are not included in this manuscript.

